# Gastroesophageal Malignancies Are Associated with Poorer Outcomes in Young Patients Seen in California Comprehensive Cancer Centers

**DOI:** 10.1101/2024.11.11.24317130

**Authors:** Armon Azizi, Aditya Mahadevan, Elaine Chiao, Maheswari Senthil, Farshid Dayyani

## Abstract

While the overall incidence of adenocarcinomas of the stomach and esophagus (GEA) has been declining in the United States, there has been an increase in cases among those under 50. Clinical and demographic data on GEA in young patients, especially in the United States, is limited. A retrospective cohort study was performed using the TrinetX database to investigate the association between patient demographics, geography, and clinical outcomes in patients with GEA between academic centers in California and a national, multi-institutional cohort. Young patients in both cohorts were more likely to be female, Hispanic, and have metastatic disease at the time of diagnosis. In California, young patients with GEA had significantly worse overall survival compared to older patients (HR 0.69, p = 0.002) whereas nationally, young patients did significantly better than old patients (HR 1.4, p < 0.0001). These disparate survival outcomes among the young UC cohort raises significant concerns and warrants further investigation.

## Introduction

Adenocarcinomas of the stomach and the esophagus (GEA) combined are the 5^th^ most common malignancy diagnosed globally.^1^ While the overall incidence of GEA in individuals over 50 has been steadily declining in the United States since the early 2000s, there has been an increase in cases among younger patients.^2–5^ This trend is part of a broader rise in early-onset cancers, with gastrointestinal cancers showing the fastest-growing incidence.^4^ In the US, GEA remain a leading cause of cancer-related death, with gastric and esophageal five-year survival rates of 21% and 36% respectively.^5^ Notably, in the United States, young adults diagnosed with GEA are more likely to present with metastatic disease compared to older adults, a concerning trend given the 30% increase in incidence among those under 50 between 2000 and 2021.^3,5,9^

To date, clinical and demographic data on GEA in young patients, especially in the United States, is limited. Some literature suggests that while young patients with non-metastatic gastroesophageal cancer tend to have better clinical outcomes than older patients, the presence of metastatic disease portends poorer prognoses in these patients.^11–13^ Additionally, the incidence of gastrointestinal malignancies in young Hispanic males is growing disproportionately.^4,14^ This is especially of significance given that clinical factors, treatment modalities, and outpatient support have been shown to differ by race in the treatment of young patients with cancer.^15,16^ Lastly, region may play a role in GEA presentation and outcomes, with prior studies demonstrating higher rates of gastric cancer in younger individuals in the northeastern US.^9^ Considering the greater loss of life-years in younger patients, further research is needed to delineate gastric cancer outcomes and decrease mortality in this population.

While prior works have evaluated the association between age and incidence of gastric cancer, to date, region-specific associations between age and survival in GEA have yet to be evaluated. Given the diversity of California’s patient population, this analysis aims to investigate the association between patient demographics, geography, and clinical outcomes in patients with GEA between academic centers in California and a national, multi-institutional cohort. We hypothesized that there would exist geography-specific differences in outcomes between young and old patients given the differences in demographics between these regions.

## Methods

A retrospective cohort study was performed using the TrinetX database, a global research network with electronic medical record data from more than 200 million patients. Specifically, the TrinetX UC and US Collaborative Networks were utilized which comprise of University of California Health Care Organizations (HCOs) or National HCOs. Using these two networks, patients were identified based on the presence of ICD codes associated with gastric and esophageal cancers (C16 & C15 respectively). Subsequently, the cohorts were partitioned by age (>= 50 and < 50). Other variables collected included race, ethnicity, gender, presence of metastatic disease (defined by ICD C79), and the occurrence of surgical intervention.

For survival analyses, young patients were matched using sex, race, ethnicity, and presence of metastases to old patients using 1:1 propensity score matching. Kaplan Meier, survival hazard ratio, and significance were then calculated with an index event of gastric/esophageal cancer diagnosis. Mortality was determined based on the time from index event to recorded death. Hazard ratios were calculated to determine the association between age group and mortality. Institutional Review Board approval was exempt for this study, as the TriNetX data adheres to the HIPAA Privacy Rule’s deidentification standard. A p value threshold of 0.05 was used for determining significance of statistical tests. All statistical analyses were performed on the TrinetX platform.

## Results

After inclusion/exclusion, the UC cohort consisted of 5590 patients and the national cohort consisted of 143,908 patients. When numerically comparing gastric and esophageal cancer patients between the UC and National cohorts, patients within the UC network had a younger age at diagnosis (Mean 64.7 vs 65.7). UC patients were more likely to be female (34.17% vs 30.55%), Hispanic (11.99% vs 6.4%), Black/African American (17.89% vs 10.13%), and Asian (9.12% vs 5%). UC patients were less likely to be white (51.7% vs 63.03%) (Table 1).

**Table 1:** Demographic Data for UC and National Cohorts.

|  | UC Cohort | National Cohort |
| --- | --- | --- |
|  | n=5590 | n=143908 |
| Age at Diagnosis (Mean ± SD) | 71 ± 13 | 74 ± 13 |
| Sex |  |  |
| Male | 3680 (65.83%) | 95080 (66.07%) |
| Female | 1910 (34.17%) | 43964 (30.55%) |
| Unknown | 10 (0.18%) | 4864 (3.38%) |
| Ethnicity |  |  |
| Not Hispanic or Latino | 2920 (52.23%) | 85136 (59.16%) |
| Unknown Ethnicity | 2010 (35.96%) | 49562 (34.44%) |
| Hispanic or Latino | 670 (11.99%) | 9210 (6.4%) |
| Race |  |  |
| White | 2890 (51.7%) | 90705 (63.03%) |
| Unknown Race | 1020 (18.25%) | 25241 (17.54%) |
| Black or African American | 1000 (17.89%) | 14578 (10.13%) |
| Asian | 510 (9.12%) | 7195 (5%) |
| Other Race | 150 (2.68%) | 4591 (3.19%) |
| Native Hawaiian or Other Pacific Islander | 30 (0.54%) | 1108 (0.77%) |
| American Indian or Alaska Native | 20 (0.36%) | 489 (0.34%) |

When comparing young (less than 50) and old (greater than or equal to 50) patients with GEA within the UC cohort, young patients were more likely to be female (48% vs 33%, std diff = 0.31, p < 0.0001) and Hispanic (24% vs 11%, std diff = 0.34, p < 0.0001). Old patients were more likely to be white (53% vs 37%, std diff = 0.32, p < 0.0001). Young patients in the UC cohort were more likely to have metastatic disease at diagnosis (11% vs 6%, std diff 0.19, p < 0.0001). In the national cohort, young patients were more likely to be female (43% vs 30%, std diff 0.28, p < 0.0001) and Hispanic (19% vs 6%, std diff = 0.38, p < 0.0001) (Table 2). Young patients in the National cohort were more likely to have metastatic disease at diagnosis (11% vs 7%, std diff 0.15, p < 0.0001).

**Table 2:** Demographic differences between young and old gastroesophageal cancer patients in the UC and National Cohorts.

|  | UC Cohort |  |  |  |  | National Cohort |  |  |  |
| --- | --- | --- | --- | --- | --- | --- | --- | --- | --- |
|  | Young (<50) | Old (>50) | Std Diff | p-value |  | Young (<50) | Old (>50) | Std Diff | p-value |
|  | n = 460 | n = 5190 |  |  |  | n = 6969 | n = 129379 |  |  |
| Age at Diagnosis (Mean ± SD) | 37.5 ± 6.95 | 66.9 ± 10.9 | 3.2143 | <0.0001 | | 36 ± 9.15 | 67.2 ± 10.9 | 3.098 | <0.0001 |
| Gender |  |  |  |  |  |  |  |  |  |
| Male | 240 (52%) | 3470 (67%) | 0.3054 | < 0.0001 |  | 3690 (54%) | 78158 (66%) | 0.2492 | < 0.0001 |
| Female | 220 (48%) | 1710 (33%) | 0.3054 | < 0.0001 |  | 2964 (43%) | 35401 (30%) | 0.2813 | < 0.0001 |
| Unknown Gender | 10 (2%) | 10 (0%) | 0.1839 | < 0.0001 |  | 182 (3%) | 4697 (4%) | 0.0732 | < 0.0001 |
| Ethnicity |  |  |  |  |  |  |  |  |  |
| Not Hispanic or Latino | 110 (24%) | 1920 (37%) | 0.2887 | < 0.0001 |  | 3980 (58%) | 75534 (64%) | 0.1161 | < 0.0001 |
| Hispanic or Latino | 110 (24%) | 570 (11%) | 0.3451 | < 0.0001 |  | 1284 (19%) | 7626 (6%) | 0.3781 | < 0.0001 |
| Unknown Ethnicity | 240 (52%) | 2700 (52%) | 0.001 | 0.9835 |  | 1572 (23%) | 35096 (30%) | 0.1521 | < 0.0001 |
| Race |  |  |  |  |  |  |  |  |  |
| White | 170 (37%) | 2740 (53%) | 0.3246 | < 0.0001 |  | 3802 (56%) | 77197 (65%) | 0.1986 | < 0.0001 |
| Asian | 80 (17%) | 940 (18%) | 0.0198 | 0.6866 |  | 435 (6%) | 6392 (5%) | 0.0407 | 0.0007 |
| Other Race | 130 (28%) | 910 (18%) | 0.2565 | < 0.0001 |  | 508 (7%) | 4306 (4%) | 0.1663 | < 0.0001 |
| Unknown Race | 60 (13%) | 460 (9%) | 0.1336 | 0.0031 |  | 1075 (16%) | 17126 (14%) | 0.0347 | 0.0046 |
| Black or African American | 20 (4%) | 130 (3%) | 0.1011 | 0.0189 |  | 901 (13%) | 11769 (10%) | 0.1011 | < 0.0001 |
| American Indian or Alaska Native | 10 (2%) | 20 (0%) | 0.1595 | < 0.0001 |  | 53 (1%) | 437 (0%) | 0.0538 | < 0.0001 |
| Native Hawaiian or Other Pacific Islander | 10 (2%) | 20 (0%) | 0.1595 | < 0.0001 |  | 62 (1%) | 1029 (1%) | 0.0039 | 0.7502 |
| Metastatic disease at diagnosis | 50 (11%) | 290 (6%) | 0.1926 | < 0.0001 |  | 754 (11%) | 8102 (7%) | 0.1465 | < 0.0001 |
| Procedures |  |  |  |  |  |  |  |  |  |
| Surgical Intervention | 10 (2%) | 20 (0%) | 0.1595 | < 0.0001 |  | 35 (1%) | 290 (0%) | 0.0434 | < 0.0001 |

We evaluated the overall survival of young and old patients in the UC and national cohorts. Survival analyses were performed following propensity score matching between groups using sex, race, ethnicity, and presence of metastases. In the national cohort, old patients had significantly shorter overall survival from diagnosis compared to young patients (median survival 1769 days vs not reached, HR 1.4 [1.33 - 1.50], log-rank p < 0.0001) (Figure 1). In contrast, in the UC cohort, old patients had significantly longer overall survival from diagnosis compared to young patients (median survival 2794 vs 828 days, HR 0.69 [0.54 - 0.88], log-rank p = 0.002) (Figure 1).

**Figure 1:**
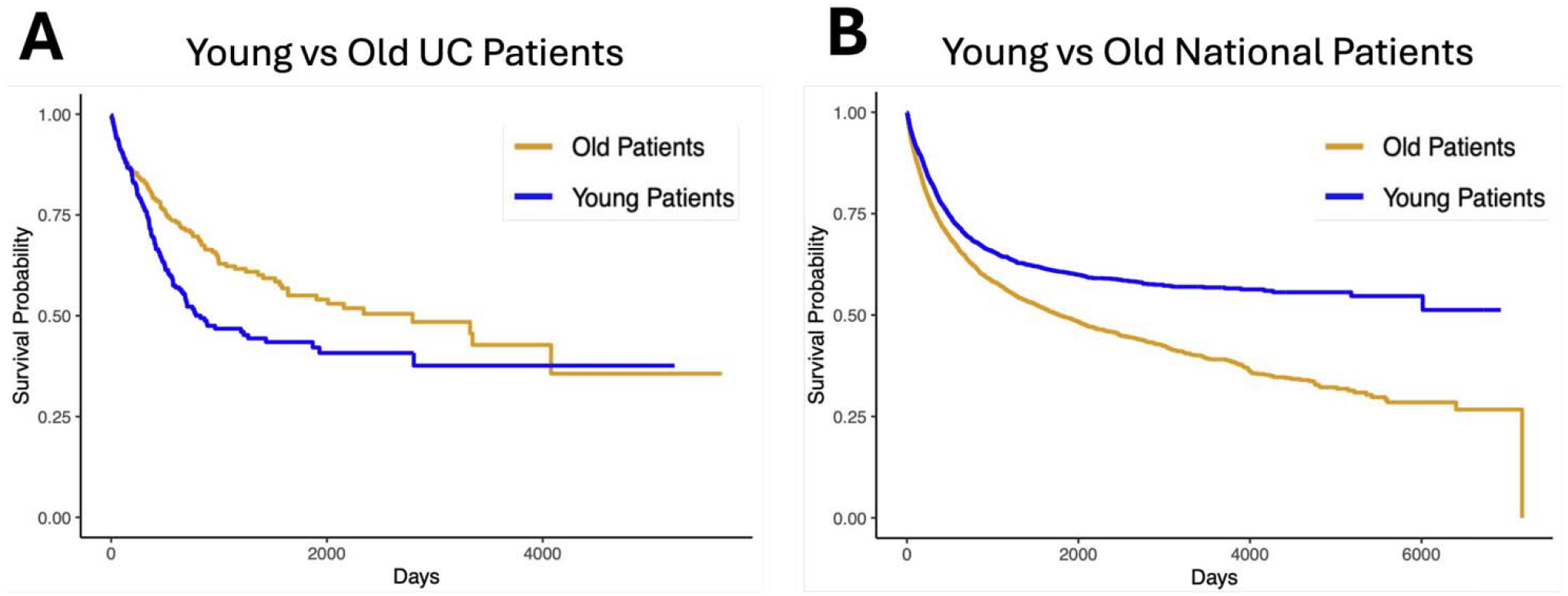
Overall survival for young vs old gastroesophageal cancer patients in the UC and National cohorts. A) Overall survival for young (<50) vs old (>50) gastroesophageal cancer patients in the University of California cohort corrected for sex, race, and metastatic status. B) Overall survival for young (<50) vs old (>50) gastroesophageal cancer patients in the National cohort corrected for sex, race, and presence of metastatic disease.

## Discussion

This analysis is the first to investigate region-specific associations between clinical and demographic characteristics and survival in patients with GEA between academic centers in California and a national, multi-institutional cohort. We show that in California, young patients with GEA have significantly worse overall survival compared to older patients whereas nationally, young patients did significantly better than old patients.

Among patients with GEA, we found some differences in demographic characteristics between the overall UC and national cohorts. UC patients were less likely to be white and more likely to be Hispanic, Black/African American, and Asian. UC patients were slightly younger at the time of diagnosis compared to the national cohort. These demographic differences are consistent with prior literature and reflective of the diversity of California’s patient population.^17,18^ Notably, in both the national and UC cohorts, young patients with GEA were more likely to be female and Hispanic, consistent with prior works demonstrating these trends in young gastric cancer patients nationally.^13,19^

Despite similar demographic trends in young gastroesophageal cancer patients in the UC and national cohorts, we found that young patients in the UC cohort had significantly worse overall survival compared to older patients whereas nationally, young patients demonstrated better overall survival compared to older patients with GEA. We note that young patients in California were more likely to be diagnosed with metastatic disease, indicating that on average, young patients in California may be diagnosed at later stages compared to young patients nationally. These differences in stage at diagnosis may, in part, explain the survival trends observed. Indeed, prior studies have suggested that stage at diagnosis and not age itself, is the primary feature that drives survival in gastric cancer.^9^

Prior works have demonstrated that young patients with gastric cancer are more likely to present with metastatic disease, however, regional differences in survival trends among young gastroesophageal cancer patients have not previously been identified. ^13^ Presentation with later-stage gastric cancer may be due to diagnostic delays, often associated with disparities in access to care and health literacy.^20,21^ Additionally, the UC cohort consisted of a higher percentage of ethnic minorities, groups previously associated with barriers to cancer screening and diagnosis.^22–24^ Taken together, this data suggests that barriers to care or screening delays for young patients in California, partially driven by demographic or socioeconomic differences, may result in poorer outcomes in young patients with GEA.

Young gastroesophageal cancer patients in California had worse outcomes than older patients, even when correcting for sex, race/ethnicity, and presence of metastases at diagnosis, suggesting that factors other than disease stage affected outcomes in these patients. Aggressive cancer subtypes such as signet cell adenocarcinoma may contribute to poorer patient outcomes in this population.^19,25,26^ Signet cell adenocarcinoma is particularly common among young, Hispanic patients, well represented in our UC cohort, further strengthening this potential link.^27^ Additionally, genomic differences, such as mutations in genes like CDH1, may play a role in geography-specific clinical outcomes.^28^ While we hypothesized that race was a significant contributing factor, the persistence of these trends despite correcting for race and ethnicity suggests that these factors are less likely to contribute to outcomes in younger patients with GEA.^29^

This study’s limitations include reliance on retrospective data, potential inaccuracies in the recording of ICD-10 codes, and the absence of detailed staging data or pathological information. Nevertheless, our analysis highlights troubling trends in GEA outcomes among younger patients in California. In particular, the higher incidence of metastatic disease raises concerns about delays in diagnosis and the potential barriers that may exacerbate adverse outcomes in this population. Further research is needed to characterize underlying factors driving poor outcomes in young patients with GEA.

## Data Availability

This retrospective study was performed using the TrinetX database. Therefore, no new data were generated or analyzed in this manuscript.

## References

1. Bray F, Laversanne M, Sung H, et al. Global cancer statistics 2022: GLOBOCAN estimates of incidence and mortality worldwide for 36 cancers in 185 countries. CA Cancer J Clin. 2024;74(3):229–263. doi:10.3322/caac.21834

2. Rodriguez GM, DePuy D, Aljehani M, et al. Trends in Epidemiology of Esophageal Cancer in the US, 1975-2018. JAMA Netw Open. 2023;6(8):e2329497. doi:10.1001/jamanetworkopen.2023.29497

3. Codipilly DC, Sawas T, Dhaliwal L, et al. Epidemiology and Outcomes of Young Onset Esophageal Adenocarcinoma: An Analysis from a Population-Based Database. Cancer Epidemiol Biomark Prev Publ Am Assoc Cancer Res Cosponsored Am Soc Prev Oncol. 2021;30(1):142–149. doi:10.1158/1055-9965.EPI-20-0944

4. Koh B, Tan DJH, Ng CH, et al. Patterns in Cancer Incidence Among People Younger Than 50 Years in the US, 2010 to 2019. JAMA Netw Open. 2023;6(8):e2328171. doi:10.1001/jamanetworkopen.2023.28171

5. National Cancer Institute. Surveillance, Epidemiology, and End Results Program. Standard populations (millions) for age-adjustment. Accessed September 9, 2024. https://seer.cancer.gov/statistics-network/explorer/application.html

6. Uhlenhopp DJ, Then EO, Sunkara T, Gaduputi V. Epidemiology of esophageal cancer: update in global trends, etiology and risk factors. Clin J Gastroenterol. 2020;13(6):1010–1021. doi:10.1007/s12328-020-01237-x

7. Smyth EC, Nilsson M, Grabsch HI, Grieken NC van, Lordick F. Gastric cancer. The Lancet. 2020;396(10251):635–648. doi:10.1016/S0140-6736(20)31288-5

8. Machlowska J, Baj J, Sitarz M, Maciejewski R, Sitarz R. Gastric Cancer: Epidemiology, Risk Factors, Classification, Genomic Characteristics and Treatment Strategies. Int J Mol Sci. 2020;21(11):4012. doi:10.3390/ijms21114012

9. De B, Rhome R, Jairam V, et al. Gastric adenocarcinoma in young adult patients: patterns of care and survival in the United States. Gastric Cancer Off J Int Gastric Cancer Assoc Jpn Gastric Cancer Assoc. 2018;21(6):889–899. doi:10.1007/s10120-018-0826-x

10. Li J. Gastric Cancer in Young Adults: A Different Clinical Entity from Carcinogenesis to Prognosis. Gastroenterol Res Pract. 2020;2020:9512707. doi:10.1155/2020/9512707

11. Puhr HC, Karner A, Taghizadeh H, et al. Clinical characteristics and comparison of the outcome in young versus older patients with upper gastrointestinal carcinoma. J Cancer Res Clin Oncol. 2020;146(12):3313–3322. doi:10.1007/s00432-020-03302-x

12. Cormedi MCV, Katayama MLH, Guindalini RSC, Faraj SF, Folgueira MAAK. Survival and prognosis of young adults with gastric cancer. Clinics. 2018;73(Suppl 1):e651s. doi:10.6061/clinics/2018/e651s

13. Calderillo-Ruíz G, Díaz-Romero MC, Carbajal-López B, et al. Latin American young patients with gastric adenocarcinoma: worst prognosis and outcomes. J Gastrointest Oncol. 2023;14(5):2018–2027. doi:10.21037/jgo-23-259

14. Merchant SJ, Kim J, Choi AH, Sun V, Chao J, Nelson R. A Rising Trend in the Incidence of Advanced Gastric Cancer in Young Hispanic Men. Gastric Cancer Off J Int Gastric Cancer Assoc Jpn Gastric Cancer Assoc. 2017;20(2):226–234. doi:10.1007/s10120-016-0603-7

15. LaPelusa M, Shen C, Gillaspie EA, et al. Variation in Treatment Patterns of Patients with Early-Onset Gastric Cancer. Cancers. 2022;14(15):3633. doi:10.3390/cancers14153633

16. Vitiello GA, Hani L, Wang A, et al. Clinical Presentation Patterns and Survival Outcomes of Hispanic Patients with Gastric Cancer. J Surg Res. 2021;268:606–615. doi:10.1016/j.jss.2021.07.031

17. Lee E, Tsai KY, Zhang J, et al. Population-based evaluation of disparities in stomach cancer by nativity among Asian and Hispanic populations in California, 2011-2015. Cancer. 2024;130(7):1092–1100. doi:10.1002/cncr.35141

18. E D, L D, Bu W. Racial and Ethnic Minorities at Increased Risk for Gastric Cancer in a Regional US Population Study. Clin Gastroenterol Hepatol Off Clin Pract J Am Gastroenterol Assoc. 2017;15(4). doi:10.1016/j.cgh.2016.11.033

19. Holowatyj AN, Ulrich CM, Lewis MA. Racial/ethnic patterns of young-onset noncardia gastric cancer. Cancer Prev Res Phila Pa. 2019;12(11):771–780. doi:10.1158/1940-6207.CAPR-19-0200

20. Ju MR, Alterio RE, Sawas T, Zeh HJ, Wang SC, Porembka MR. Gaps in Providers’ Knowledge Delays Gastric Cancer Diagnosis. J Gastrointest Surg. 2022;26(4):750–756. doi:10.1007/s11605-021-05209-5

21. Guadamuz JS, Wang X, Ryals CA, et al. Socioeconomic status and inequities in treatment initiation and survival among patients with cancer, 2011-2022. JNCI Cancer Spectr. 2023;7(5):pkad058. doi:10.1093/jncics/pkad058

22. Vrinten C, Gallagher A, Waller J, Marlow LAV. Cancer stigma and cancer screening attendance: a population based survey in England. BMC Cancer. 2019;19. doi:10.1186/s12885-019-5787-x

23. Shokar NK, Vernon SW, Weller SC. Cancer and colorectal cancer: knowledge, beliefs, and screening preferences of a diverse patient population. Fam Med. 2005;37(5):341–347.

24. Gh R, Ce F, K K, Rt C, Ee C, Rb W. Misconceptions about breast lumps and delayed medical presentation in urban breast cancer patients. Cancer Epidemiol Biomark Prev Publ Am Assoc Cancer Res Cosponsored Am Soc Prev Oncol. 2010;19(3). doi:10.1158/1055-9965.EPI-09-0997

25. Graziosi L, Marino E, Natalizi N, Donini A. Prognostic Survival Significance of Signet Ring Cell (SRC) Gastric Cancer: Retrospective Analysis from a Single Western Center. J Pers Med. 2023;13(7):1157. doi:10.3390/jpm13071157

26. Choi AH, Ji L, Babcock B, et al. Peritoneal carcinomatosis in gastric cancer: Are Hispanics at higher risk? J Surg Oncol. 2020;122(8):1624–1629. doi:10.1002/jso.26210

27. Yao JC, Tseng JF, Worah S, et al. Clinicopathologic behavior of gastric adenocarcinoma in Hispanic patients: analysis of a single institution’s experience over 15 years. J Clin Oncol Off J Am Soc Clin Oncol. 2005;23(13):3094–3103. doi:10.1200/JCO.2005.08.987

28. Benesch MGK, Bursey SR, O’Connell AC, et al. CDH1 Gene Mutation Hereditary Diffuse Gastric Cancer Outcomes: Analysis of a Large Cohort, Systematic Review of Endoscopic Surveillance, and Secondary Cancer Risk Postulation. Cancers. 2021;13(11):2622. doi:10.3390/cancers13112622

29. Laszkowska M, Tramontano AC, Kim J, et al. Racial and ethnic disparities in mortality from gastric and esophageal adenocarcinoma. Cancer Med. 2020;9(15):5678–5686. doi:10.1002/cam4.3063

30. Ajani JA, D’Amico TA, Bentrem DJ, et al. Gastric Cancer, Version 2.2022, NCCN Clinical Practice Guidelines in Oncology. J Natl Compr Cancer Netw JNCCN. 2022;20(2):167–192. doi:10.6004/jnccn.2022.0008

